# Wastewater surveillance for *Salmonella* Typhi and its association with seroincidence of enteric fever in Vellore, India

**DOI:** 10.1101/2024.07.25.24310996

**Authors:** Dilip Abraham, Lalithambigai Kathiresan, Midhun Sasikumar, Kristen Aiemjoy, Richelle C. Charles, Dilesh Kumar, Rajan Srinivasan, Catherine Troman, Elizabeth Gray, Christopher B. Uzzell, Jacob John, Balaji Veeraraghavan, Nicholas C. Grassly, Venkata Raghava Mohan

## Abstract

**Background:** Blood culture-based surveillance for typhoid fever has limited sensitivity, encountering operational challenges in resource-limited settings. Environmental surveillance targeting *Salmonella* Typhi (*S*. Typhi) shed in wastewater (WW), coupled with cross-sectional serosurveys of *S.* Typhi-specific antibodies estimating exposure to infection, emerges as a promising alternative.

**Methods:** We assessed the feasibility and effectiveness of wastewater (WW) and sero-surveillance for S. Typhi in Vellore, India, from May 2022 to April 2023. Monthly samples were collected from 40 sites in open drainage channels and processed using standardized protocols. DNA was extracted and analyzed via quantitative PCR for S. Typhi genes (*ttr, tviB, staG*) and the fecal biomarker HF183. Clinical cases of enteric fever were recorded from four major hospitals, and a cross-sectional serosurvey measured hemolysin E (HlyE) IgG levels in children under 15 years to estimate seroincidence.

**Results:** 7.50% (39/520) of grab and 15.28% (79/517) Moore swabs were positive for all 3 *S .* Typhi genes. Moore swab positivity was significantly associated with HF183 (adjusted odds ratio (aOR): 3.08, 95% CI: 1.59 – 5.95) and upstream catchment population (aOR: 4.67, 1.97 – 11.04), and there was increased detection during monsoon season - membrane filtration (aOR: 2.99, 1.06 – 8.49), and Moore swab samples (aOR: 1.29, 0.60 – 2.79). Only 11 blood culture-confirmed typhoid cases were documented over the study period. Estimated seroincidence was 10.4/100 person-years (py) (95% CI: 9.61 - 11.5/100 py). The number of *S.* Typhi positive samples at a site was associated with the estimated sero- incidence in the site catchment population (incidence rate ratios: 1.14 (1.07-1.23) and 1.10 (1.02-1.20) for grab and Moore swabs respectively.

**Conclusions:** These findings underscore the utility and effectiveness of alternate surveillance approaches to estimating the incidence of S. Typhi infection in resource-limited settings, offering valuable insights for public health interventions and disease monitoring strategies where conventional methods are challenging to implement.

**Author Summary:** Our study explores the potential of detecting *Salmonella* Typhi in wastewater, informing public health strategies, guiding vaccination campaigns, and monitoring the effectiveness of interventions, contributing to better disease control and prevention policies.

*S*. Typhi positivity rates of 7.50% and 15.30% were observed in grab samples and Moore swabs, respectively, and significant correlations were found between *S*. Typhi positivity in Moore swabs and fecal contamination marker levels in wastewater samples.

The study also estimated that the community seroincidence for typhoid 10.40/100 person-years and *S*. Typhi detections in WW positively correlated with seroincidence.

The study reveals significant associations between wastewater S. Typhi positivity and typhoid seroincidence, seasonal variations, and population dynamics, providing deeper insights into the epidemiology of typhoid fever.

## Introduction

Globally, approximately 11 million new cases of typhoid infection occur annually, resulting in 77,000 to 219,000 deaths [1], mainly driven by the high incidences in Southeast Asia and sub-Saharan Africa, with India being one of the countries with the highest incidences [2]. Despite significant improvements in access to safe drinking water and sanitation in India over recent decades, typhoid remains a major challenge, especially in urban and peri-urban areas. Vellore, a city in Tamil Nadu, has a high incidence of typhoid fever. From 2017 to 2020, the annual incidence among children aged 0–14 was estimated at 1,173 cases per 100,000 child years [3,4], with a positivity rate of 3.5% for suspected enteric fevers in Tier 1 community-based surveillance, as described in the Surveillance for Enteric Fever in India (SEFI) study [4].

Typhoid incidence in urban and peri-urban Indian communities is often underestimated due to frequent early self-medication with readily available over-the-counter antibiotics, reducing blood culture sensitivity. This indiscriminate antibiotic use increases the risk of antimicrobial resistance, with the emergence of extensively drug-resistant *S*. Typhi strains [5,6]. In 2020, the World Health Organization (WHO) recommended Typhoid Conjugate Vaccines to control typhoid fever, prioritizing countries with high disease burden or record of antimicrobial-resistant *S.* Typhi [7]. However, data on typhoid fever in Low- and Middle-Income Countries (LMICs) are limited due to insufficient resources for blood culture surveillance, impeding vaccine introduction [8]. Microbiological culture, despite limitations in sensitivity, is the reference standard for *S.* Typhi detection in clinical practice and surveillance, and no other diagnostic test currently matches the accuracy of blood cultures for disease surveillance. However, it is time-consuming and resource-intensive, complicating detection in typhoid-endemic LMICs [9,10].

Environmental surveillance (ES) of sewage and wastewater (WW) samples is a cost-effective alternative to blood culture-based fever surveillance for detecting microorganisms [11]. ES can estimate subclinical disease transmission within communities. Currently, ES is mainly used in polio surveillance to complement vaccination programs, aiding in identifying transmission and guiding targeted vaccination efforts [12]. The COVID-19 pandemic has further promoted ES as a valuable tool for monitoring infectious diseases, including respiratory infections [13,14]. ES-based surveillance has identified *S.* Typhi in WW in Nepal, Pakistan, India, Malawi, and Bangladesh [11,15,16].

Potential use cases for ES of *S.* Typhi include supporting routine vaccination, monitoring intervention strategies (e.g., vaccines or improved water, sanitation, and hygiene), detecting emergence and transmission of *S.* Typhi in non-endemic regions, and identifying antimicrobial-resistant lineages [17]. Despite its potential for typhoid control, programmatic surveillance networks are scarce due to limited data linking environmental detection of *S.* Typhi with disease burden. As a host-restricted pathogen, *S.* Typhi is challenging to culture from WW, necessitating the intervention of molecular detection methods. However, regional variations in WW sample matrices lead to significant differences in molecular detection, and there is no consensus on laboratory processing methods and analytical techniques to model community disease burden [17]. Recognizing the disruption caused by COVID-19 lockdowns on healthcare-seeking behavior and surveillance measures, the study included a cross-sectional population-based serosurvey which utilized a novel serological assay based on assessing IgG responses to Hemolysin E (HlyE) antigen in plasma/serum samples to estimate seroincidence, serving as a surrogate measure of enteric fever (typhoid and paratyphoid) burden during the pandemic period [18]. This technique allows for estimating population-level seroincidence using antibody decay rates fitted to a Bayesian hierarchical model, providing insight into exposure to *S*. Typhi and *S*. Paratyphi A over the preceding year. Previous comparison of seroincidence against clinical incidence from blood culture samples across four different countries revealed that population-level seroincidence rates closely mirrored the rank order of clinical incidences. The key advantage of this method lies in its ability to estimate disease exposure within populations, even from asymptomatic cases. This approach may offer a better correlation to ES detection than diagnosing infection solely based on overt clinical cases of enteric fever. However, this method also has limitations since HlyE cannot discriminate *S*. Typhi from *S*. Paratyphi A, and reinfections may modify the antibody responses in high-exposure settings. This study sought to demonstrate the feasibility of ES as an alternative to clinical surveillance for typhoid in resource-limited settings and to compare environmental detection using grab and trap sampling techniques. We examined the association between *S.* Typhi in WW samples and clinical incidence captured through hospital-based surveillance using blood culture diagnosis, and this has been reported earlier in two study settings [16] alongside community-based seroincidence estimated via a cross-sectional survey.

## Methods

The study protocol, design, site selection, and laboratory assays have been described previously [19]. This multisite study was carried out in Vellore and Blantyre, Malawi. ES was integrated with ongoing hybrid surveillance for typhoid, including blood culture-based surveillance and a community serosurvey. This study was approved by the Institutional Review Board of Christian Medical College Vellore (IRB MIN 11170, A23 22-07-2020).

### Study area

Vellore city (12.92°N, 79.13°E), with a population of 613,000, is the administrative headquarters of the Vellore district, located on the Palar Riverbank in northeastern Tamil Nadu, India. Vellore has four zones (a total of 60 wards) that cover 87.915 km^2^ and a population of approximately 500,000 (Government of India Census, 2011). Vellore experiences a semi-arid climate with high annual temperatures and relatively low rainfall. The city has three seasons: summer (March–July, temperatures > 40°C), rains (August– November, with both southwest and northeast monsoons), and winter (December–February, low of 15°C). The average annual rainfall is 1053 mm, with approximately 60% occurring during the rainy season. The surveillance area is within the Vellore Health and Demographic Surveillance System (VHDSS) maintained by Christian Medical College, Vellore.

### ES site selection

The sewage networks in the study area consist of shallow open-drain channels that drain upstream residential areas and eventually flow north into either a Sewage Treatment Plant (STP) or directly into the dry Palar riverbed. Candidate ES sites (sampling points) were systematically identified from the southern half of the city (280,000 population) at WW and sewage confluence points using a remote GIS-based approach with environmental data coupled with a novel selection process. The site selection process involved using digital elevation models to map wastewater confluence sites to upstream catchment areas using the standardised AGREE watershed delineation approach as previously described [16,19]. Forty sampling sites were selected to represent varying catchment population sizes and densities across the study area. The field team then visited these sites to assess accessibility, safety, and the perceived appropriateness of adequate depth and flow rate (Fig 1). Catchment areas for each sampling point were mapped through ground-truthing, identifying households connected to the drains contributing to each site. The catchments varied in population size from 233 to 28,036. Some of the catchments overlapped and were nested within larger catchments— among the 40 catchment areas, 32 were independent, five included 2–3 nested catchments (level 1), and three were large catchments with multiple nested catchments.

**Fig 1.**
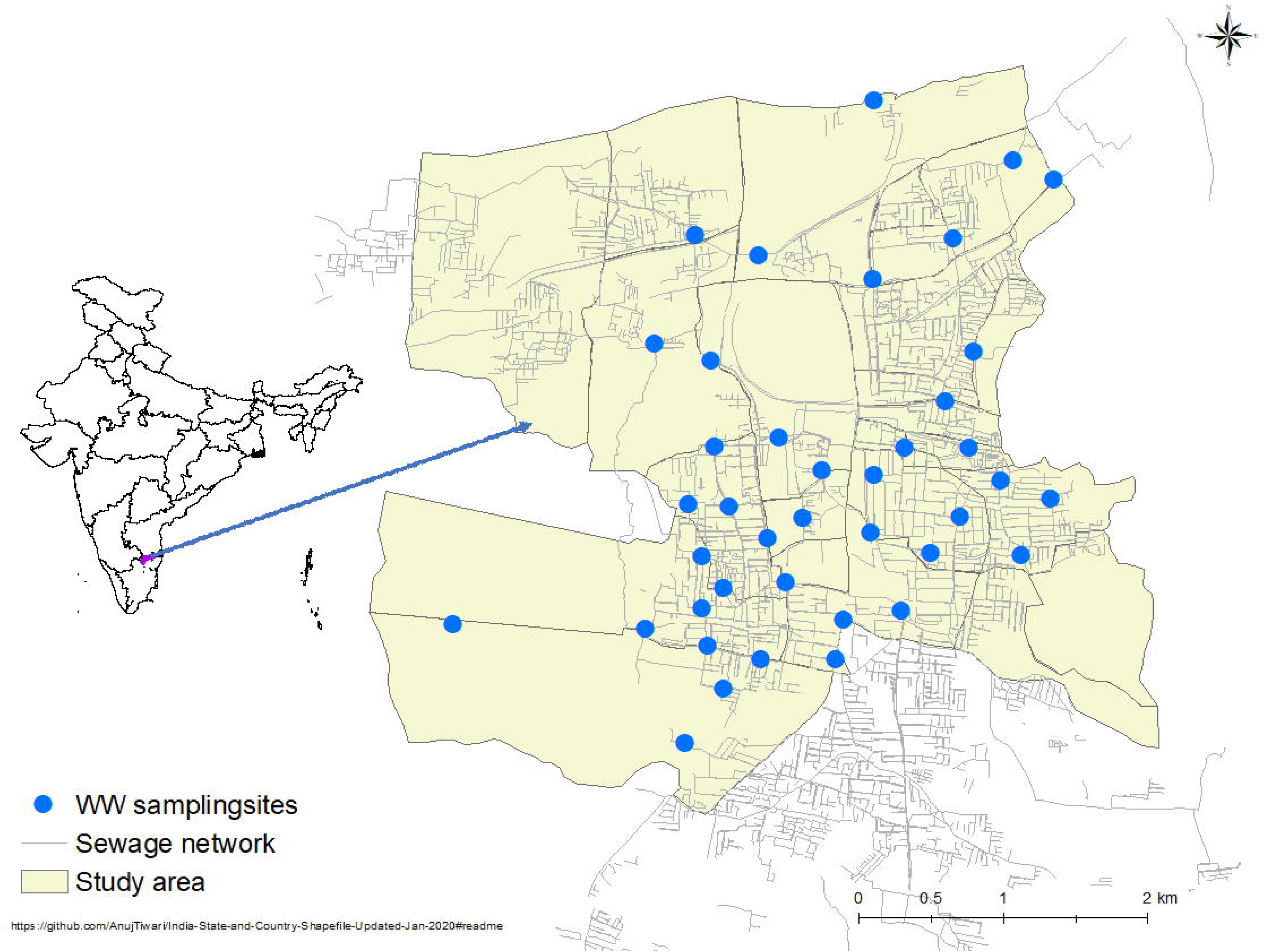
Location of the study area in Vellore, India. The yellow-shaded area in the map shows the study area in Vellore urban city with the mapped sewage network consisting of open drainage channels. The blue dots in the map represent the 40 wastewater sampling sites.

### Sample collection and processing

At each ES site, grab (membrane filtration) and trap (Moore swab) samples were collected monthly for a year starting May 2021. Pilot studies were conducted at the site to compare the recovery efficiencies of grab and trap samples, and both methods were employed for sample processing, as described in the protocol [19].

Moore swabs were prepared as described in a previous study [20] and were deployed 48 hours before collection. Grab samples were collected concurrently as the Moore swabs were deployed, with a recommended collection time between 6 am and 10 am. Moore swabs were incubated overnight in universal pre-enrichment (UPE) broth before filtration using 0.45μm membrane filters and stored at −20°C until DNA extraction. Approximately 1 L of grab samples were collected and filtered (0.45μm) after pre-filtering through a coffee filter, with filters eluted in Ringer’s lactate. The eluate was then centrifuged, and the pellet was stored at −20°C [21]. Personal protective equipment was worn during sample collection, and infection control procedures were followed to minimize risks and prevent contamination.

Physicochemical parameters, including temperature, pH, oxidative reduction potential, dissolved oxygen, total suspended solids, salinity, and turbidity were measured and recorded using water quality probes (AquaProbe AP-2000; Aquaread Ltd. UK).

### DNA extraction and PCR strategy

DNA was extracted using the QIAamp® PowerFecal® Pro DNA Kit (Qiagen), eluted into 50 mL, and stored at −20°C. Multiplex quantitative PCR (qPCR) with primers and probes targeting three genes (*ttr, staG,* and *tviB*) was used to detect *S.* Typhi. Samples in which all three targets detected were considered positive for *S.* Typhi based on their demonstrated specificity for pan-*Salmonella* species (*ttr*), *S.* Typhi, non-typhoidal *Salmonella* serovars (*staG*), or *S.* Typhi alone (*tviB*)., The cut-offs for each target were calculated as described in the published protocol paper (the Ct values used were *ttr* - 38, *staG* - 39 and *tviB* - 39) [19]. Samples positive for *ttr* and either *staG* or *tviB* alone were retested in a singleplex qPCR for the negative target to improve the sensitivity of *S.* Typhi detection in the sampling matrix [22]. Additionally, a duplex qPCR was conducted for HF183 [23], a marker gene from a human-restricted *Bacteroides*, and an extraction/PCR positive control (Cy®5-QXL®670, Eurogentec) that was spiked into samples prior to extraction. Genome copy numbers were estimated using standard curves generated using gBlocks™ DNA fragments for each target (Integrated DNA Technologies). The limit of detection (LOD) was estimated using probit regression analysis for dilutions with a 95% probability of generating a proper amplification. Laboratory assay protocols are described in detail in protocols.io (https://www.protocols.io/workspaces/typhoides).

### Clinical surveillance and estimation of incidence

The clinical surveillance data were obtained from an ongoing passive hospital blood culture-based surveillance, which collected information from patients (> 6 months of age) diagnosed with typhoid fever, referred to as ‘Tier 2’ surveillance in the SEFI study. The clinical surveillance was planned as a continuation of the Tier 2 surveillance, with further details available in the protocol of the SEFI study [4].

### Sero-survey and HlyE IgG ELISA

A cross-sectional survey was conducted on 1,200 randomly selected study participants aged between 6 months and 15 years from 20 of the 40 ES catchment areas between September 30 and November 3, 2022. Blood samples were collected after obtaining parental consent. Plasma was separated and stored at -70°C until tested for anti-HlyE IgG by ELISA as previously described [18]. Briefly, Nunc Maxisorb microplates were coated with purified HlyE (1μg/mL), and plasma samples were added at 1:5000 dilution. Bound antibodies were detected with anti-human IgG conjugated with horseradish peroxidase (Jackson ImmunoResearch) and o-phenylenediamine was used as the substrate. Kinetic readings of the plates were done at 450 nm and sample values were normalized to HlyE IgG monoclonal control included on each plate.

### Weekly average rainfall measurements, depth, and flow

Daily meteorological data on rainfall, humidity, and temperature were sourced from the Regional Meteorological Center, India Meteorological Department website for the study period [24]. WW drains were classified as ‘shallow,’ ‘medium’, or ‘deep’ if the depth of the WW levels were ‘<5 cm’, ‘5 to 50 cm’, and ‘>50 cm’, respectively. WW flow speeds were categorized into either ‘stagnant,’ ‘slow’, or ‘fast-flowing’ based on observation at the sampling time.

### Ethics statement

The Wastewater surveillance work has been approved by the Institutional Review Board of Christian Medical College, Vellore; CMC IRB Min No.11170. amended on 22 July 2020, IRB number A23 – 22.07.2020. Community serosurvey for estimating the seroincidence of Typhoid was approved by the Institutional Review Board of Christian Medical College, Vellore; CMC IRB Min No.12973 (OBSERVE) dated 24 June 2020. Formal informed consent was obtained from the parent/guardian of all children for serosurvey, and written informed assent was obtained from children aged 11 to 15 years.

### Statistical analysis

Variations in WW physicochemical parameters across sites were analyzed using analysis of variance (ANOVA). A mixed-effects logistic regression was used to determine the association between *S.* Typhi detection and WW parameters, site characteristics, weekly average rainfall, catchment population, and HF183 levels. The association of WW characteristics with Moore swab positivity was not assessed as it remains in the WW for 48 hours from deployment, and those parameters may vary between the time of deployment and collection. A random effect by location was included to account for repeated observations. Univariate analysis was used to determine the association with *S.* Typhi detection, followed by multivariate analysis for statistically significant variables (p < 0.05). Additionally, exploratory analyses of the spatial distribution of *S.* Typhi were also performed. Seroincidence for each catchment was estimated using maximum likelihood profiles for ELISA data against the fitted Bayesian hierarchical model described by Aiemjoy *et al* (2022), assuming Poisson-distributed time between incident infections [18]. The Bayesian model was adjusted to include only longitudinal antibody decay rates in children under 15 years of age, matching the sampled population. Overall and catchment-specific seroincidence rates were estimated as incidence cases per 100 person-years for the previous year. Catchment areas that surveyed a minimum of 20 participants were included for seroincidence estimation for the previous year. WW positivity for *S.* Typhi in grab and Moore swab samples was aggregated for each catchment. In addition, overall WW positivity for a sample was defined as being positive for either the grab sample or the Moore swab. Furthermore, univariate and multivariate negative binomial regression analyses were performed to estimate the association between WW positivity and seroincidence, considering overdispersion in the WW data. Negative binomial regression was adjusted for the catchment population to estimate the association of seroincidence with overall WW positivity. All analyses were performed in R version 4.3.2 [25] using the statistical packages (ggplot2, data.table, sf, MASS, lme4) [26–30]. Geospatial analysis was performed within ArcMap v10.82 [31] using spatial analysis tools.

## Results

### *S.* Typhi in WW samples

Out of the 1037 samples collected across all sampling time points, 520 were grab samples processed using the membrane filtration method and 517 using Moore swabs, with three of the Moore swabs missing. A total of 39/520 (7.50%) grab and 79/517 (15.28%) Moore swab samples tested positive for *S.* Typhi (*ttr*, *staG*, and *tviB* were all positive). *S.* Typhi was detected in the samples collected from 26/40 (65.00%) sites during the surveillance period, with an overall positivity rate of 11.40% (118/1037). The proportion of detection varied by sampling site; membrane filtration showed a median positivity of 7.69% positivity for all catchment levels with a variance of 3.20%.

A distinct spike in overall molecular detection of *S.* Typhi with both grab and Moore swab samples was observed during the monsoon months of August – November, correlating with average rainfall in Vellore (Fig 2). Rainfall during the sampling week was significantly associated with *S*. Typhi positivity for grab sample (adjusted odds ratio [aOR]: 2.56, 95% confidence interval [CI]:1.02–6.42), and Moore swab samples (aOR: 2.00, 95% CI: 1.01– 3.97). The grab sampling technique showed significantly elevated positivity in monsoon WW samples (aOR, 2.99; 95% CI, 1.03–8.49; Table 1). *S.* Typhi positivity in WW during the monsoon months (August–November) was the highest at 16.76 % (58/346), compared to 9.77% (25/256) and 8.05% (35/435) during the winter and summer months, respectively (Fig 3).

**Fig 2.**
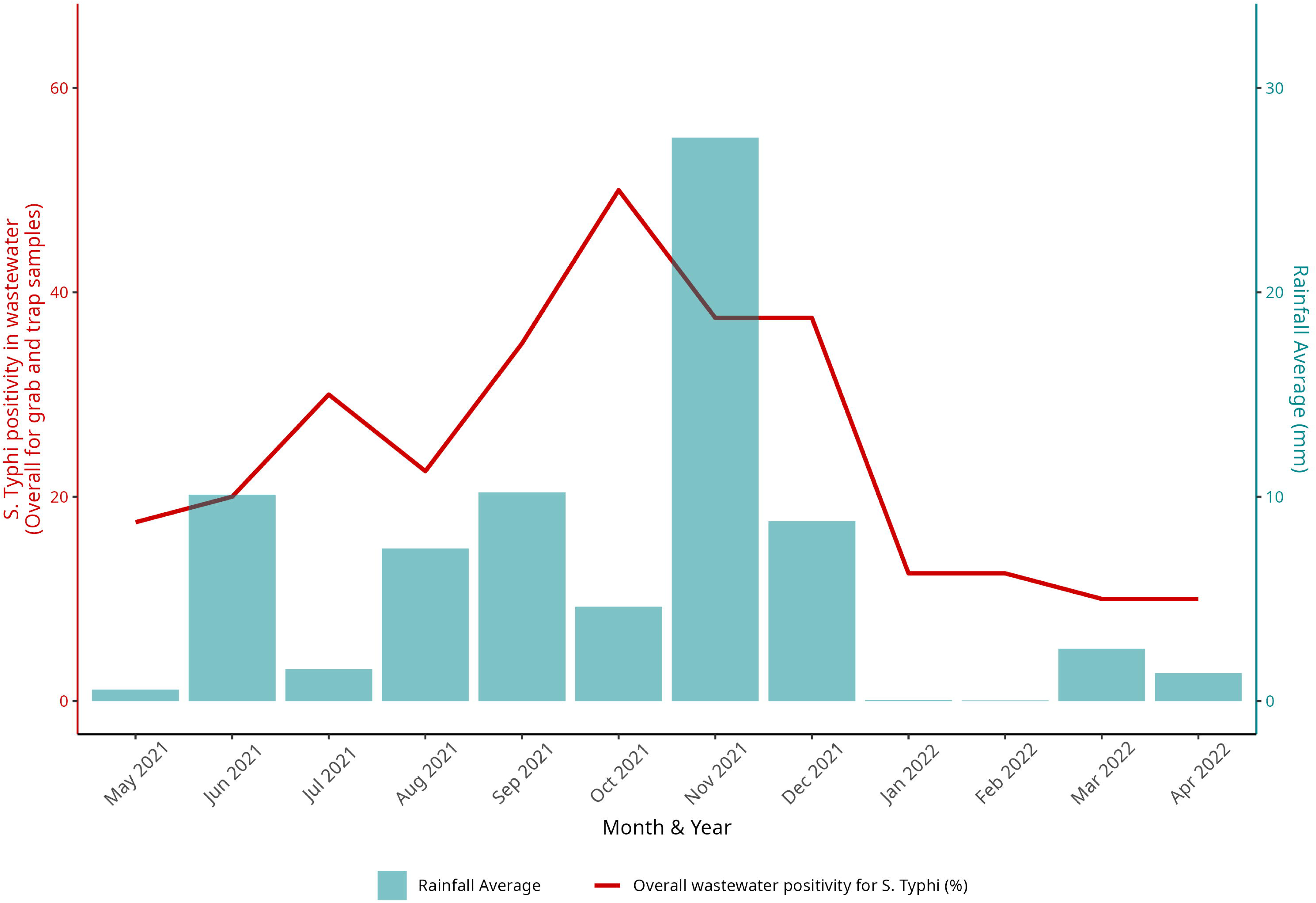
Salmonella Typhi positivity in wastewater (WW) and average monthly rainfall during the study period. The overall wastewater positivity for S. Typhi in percentage and the average monthly rainfall in millimeters is plotted over the study period from May 2021 – April 2022. The height of the bar represents the average rainfall millimeters in each month during the study period, and the line graph represents the overall positivity for S. Typhi during the same months.

**Fig 3.**
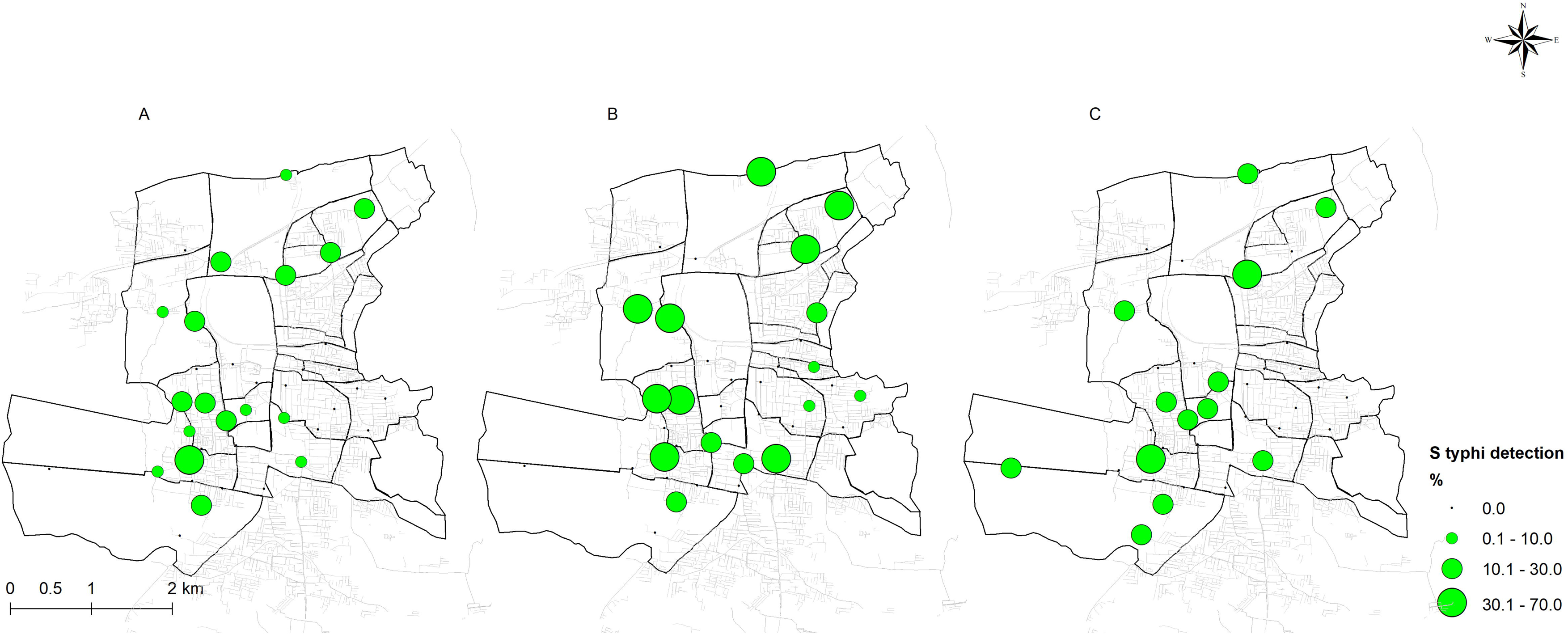
Salmonella typhi detection at wastewater sampling sites during different seasons of the year during (A) summer months (March–July), (B) monsoon months (August– November), and (C) winter months (December–February). Seasonal detection of S. Typhi detection at wastewater sampling sites is depicted in Fig 3 Panel A illustrates the positivity during the summer months (March–July), panel B during the monsoon months (August November), and panel C during the winter months (December– February). The area of the green circles represents the proportion (%) of overall detection for each period.

**Table 1.**
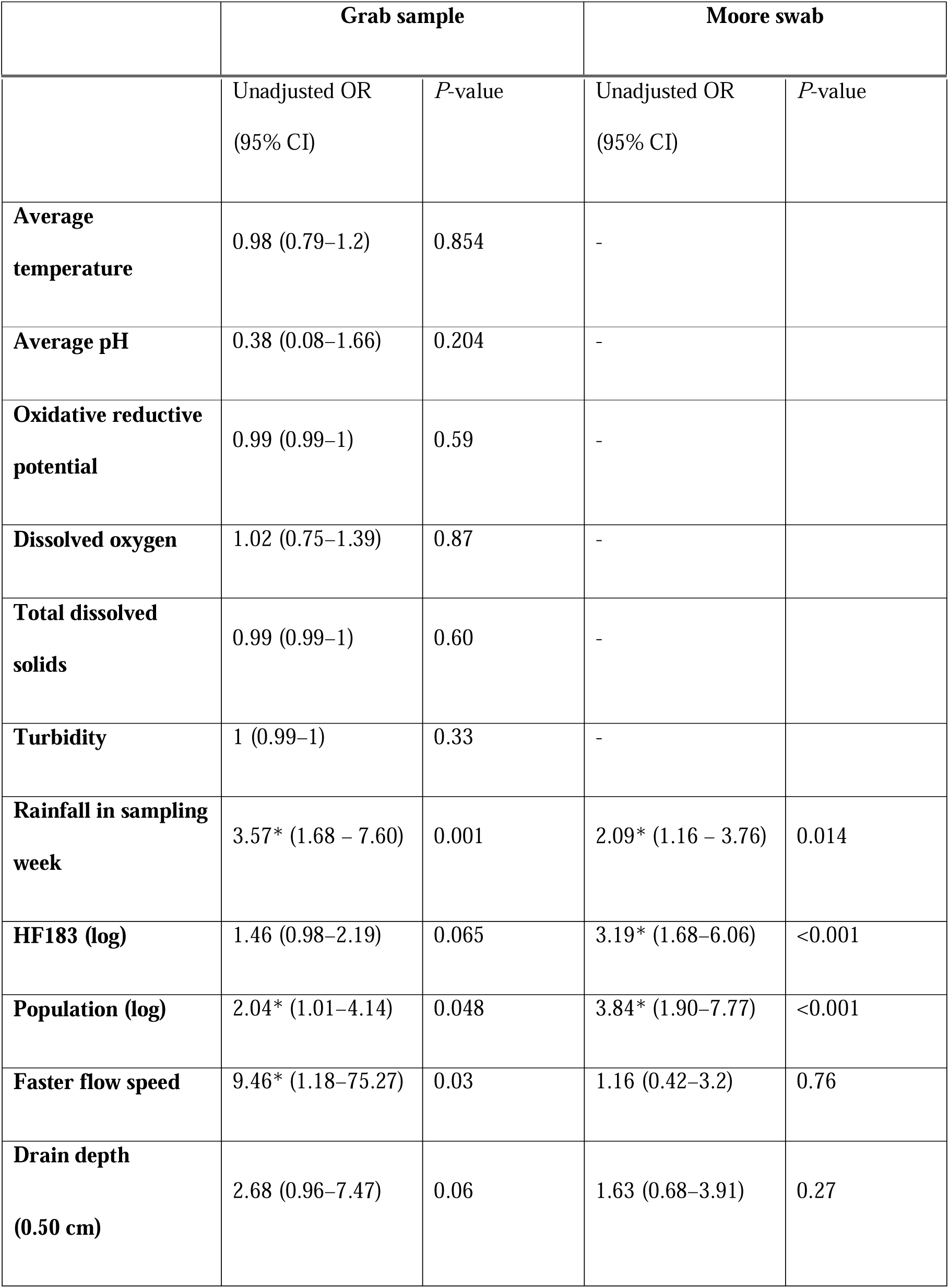
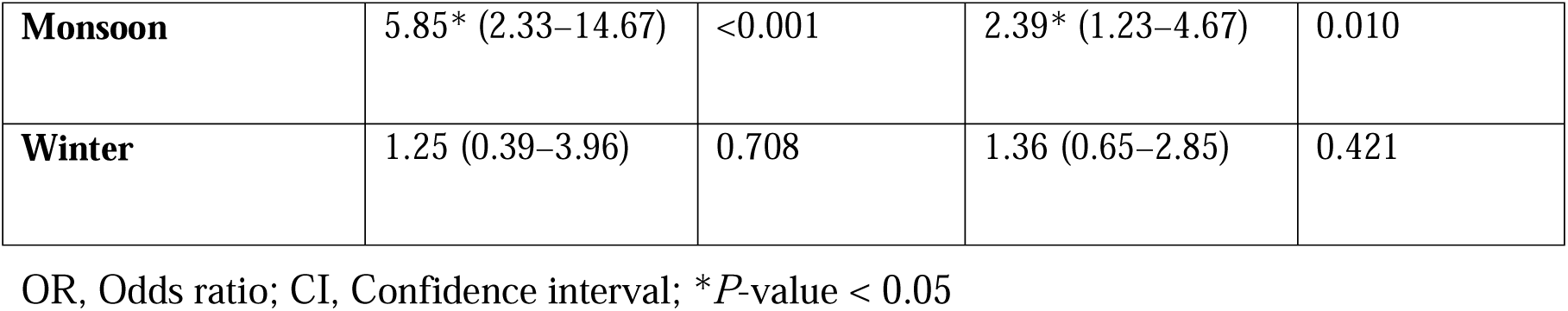
Mixed-effects logistic regression models predicting *S*. Typhi detection in ES with wastewater and catchment area characteristics. Table A: Results of unadjusted regression model predicting *S.* Typhi detection in ES with wastewater and catchment area characteristics

**Table 1B:**
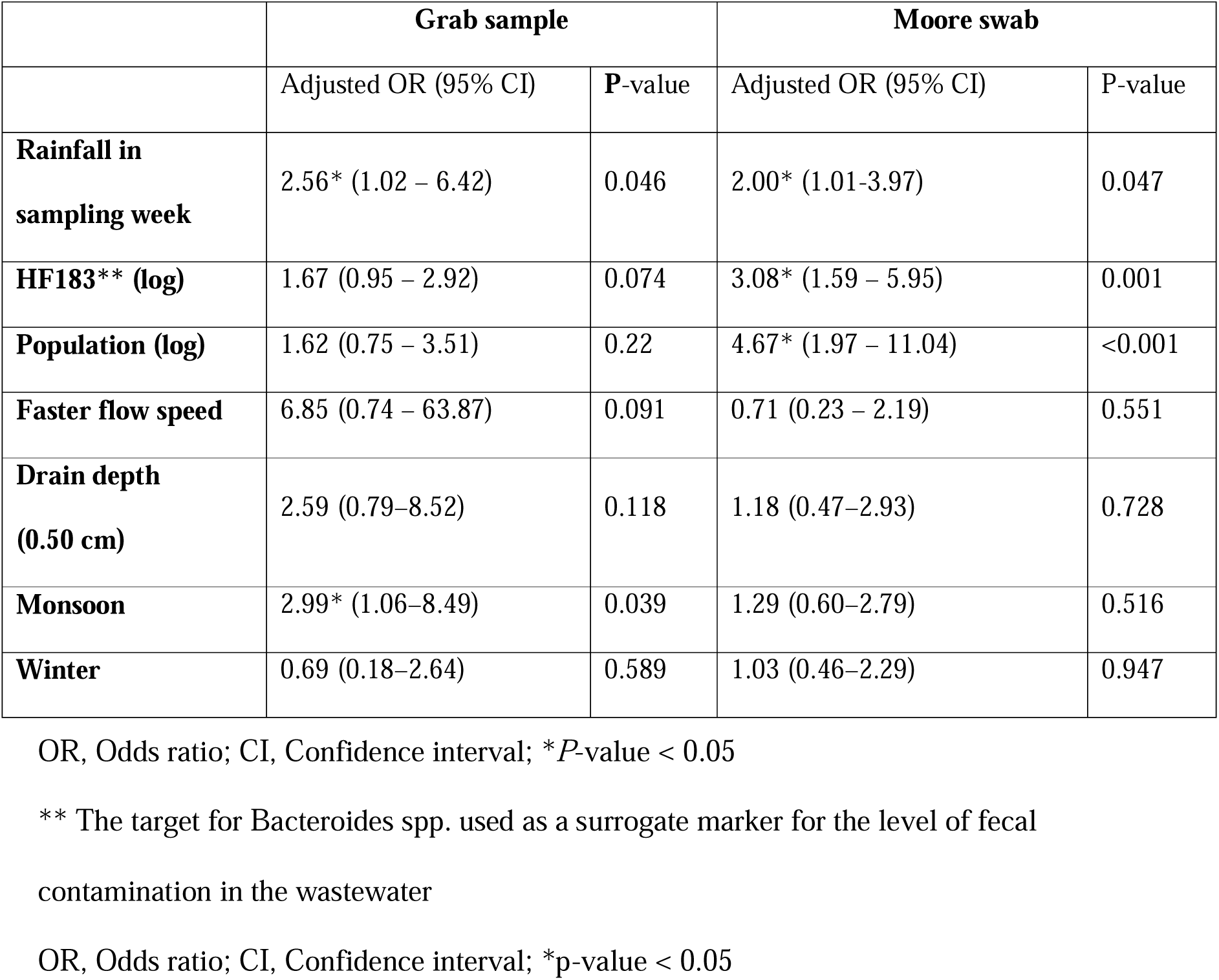
Results of adjusted regression model predicting *S.* Typhi detection in ES with wastewater and catchment area characteristics.

|  | **Grab sample** |  | **Moore swab** |  |
| --- | --- | --- | --- | --- |
|  | Adjusted OR (95% CI) | P-value | Adjusted OR (95% CI) | P-value |
| **Rainfall in sampling week** | 2.56* (1.02 – 6.42) | 0.046 | 2.00* (1.01-3.97) | 0.047 |
| **HF183\*\* (log)** | 1.67 (0.95 – 2.92) | 0.074 | 3.08* (1.59 – 5.95) | 0.001 |
| **Population (log)** | 1.62 (0.75 – 3.51) | 0.22 | 4.67* (1.97 – 11.04) | <0.001 |
| **Faster flow speed** | 6.85 (0.74 – 63.87) | 0.091 | 0.71 (0.23 – 2.19) | 0.551 |
| **Drain depth (0.50 cm)** | 2.59 (0.79–8.52) | 0.118 | 1.18 (0.47–2.93) | 0.728 |
| **Monsoon** | 2.99* (1.06–8.49) | 0.039 | 1.29 (0.60–2.79) | 0.516 |
| **Winter** | 0.69 (0.18–2.64) | 0.589 | 1.03 (0.46–2.29) | 0.947 |

OR, Odds ratio; CI, Confidence interval; \**P*-value < 0.05
\*\* The target for *Bacteroides* spp. used as a surrogate marker for the level of fecal contamination in the wastewater
OR, Odds ratio; CI, Confidence interval; \**p*-value < 0.05

### Wastewater and sampling site characteristics

All water quality parameters, except temperature, varied significantly between sites (S1 Table). In addition, a repeated-measures ANOVA revealed that all these parameters vary significantly within a site over the course of a year. Sites were characterized by flow speed during sample collection and Moore swab deployment. Of 1022 samples where flow data was available, 793/1022 (77.59%) were from fast-flowing sewage, with 107/793 (13.50%) positivity for *S*. Typhi, and 229/1022 (22.41%) from slow-flowing sewage with 10/229 (4.40%) positivity (Chi square = 10.44, p = 0.001).

Mixed-effects logistic regression showed no significant association between the WW physicochemical parameters and the detection of *S.* Typhi. Although the depth of the sewage channel and faster flow speed were significantly associated with detection in the unadjusted analysis, they did not show significant differences after adjusting for confounders in the model. Log-transformed HF183 in sewage (genome copies/μL) was associated with a significant increase in the probability of detection in Moore swab samples (aOR, 3.08; 95% CI: 1.59–5.93). Upstream catchment population size was also significantly associated with WW positivity for Moore swab samples (aOR, 4.67; 95% CI: 1.97–11.04) (Table 1). A mixed-effects linear regression indicated that HF183 copy numbers were unaffected by weekly average rainfall (*p* = 0.82) and log population (*p* = 0.95). Compared to grab samples, HF183 detection was significantly higher in Moore swab samples (OR 3.19, p < 0.01).

### Hospital cases of typhoid fever during the study period

Eleven cases of blood culture-confirmed enteric fever were documented from the study area in the sentinel hospitals during the study. Occurrence of cases was sporadic and spatial clustering was not observed as described earlier [16].

### Seroincidence estimation for ES catchments

A total of 1215 children (628 females and 587 males) were recruited for the serosurvey, of which 19.18% were aged upto two years, 21.32% were 3–5 years old, 33.00% were 6–10 years old, and 26.5% were 11–15 years old. Participants were assigned to catchment areas based on their residential locations. For seroincidence estimation, only catchment areas with a minimum of 20 participants were included, resulting in the inclusion of 1172 participants from the total 1215 surveyed.

The estimated seroincidence in this study population was 10.40/100 person-years (py) (95% CI: 9.61–11.46/100 py). Seroincidence rates varied in the non-nested, independent catchments from 6.10 to 24.60/100 py; while those in the larger catchment levels approximated the overall seroincidence estimate, ranging from 8.30 to 10.50/100 py except for one catchment that had a high estimate of 20.60/100 py. The overall wastewater positivity in these same catchments also showed similar high variability, ranging from no detections across the sampling period to a maximum of 34.61% at one site (Fig 4, S2 Table, S1 Fig).

**Fig 4.**
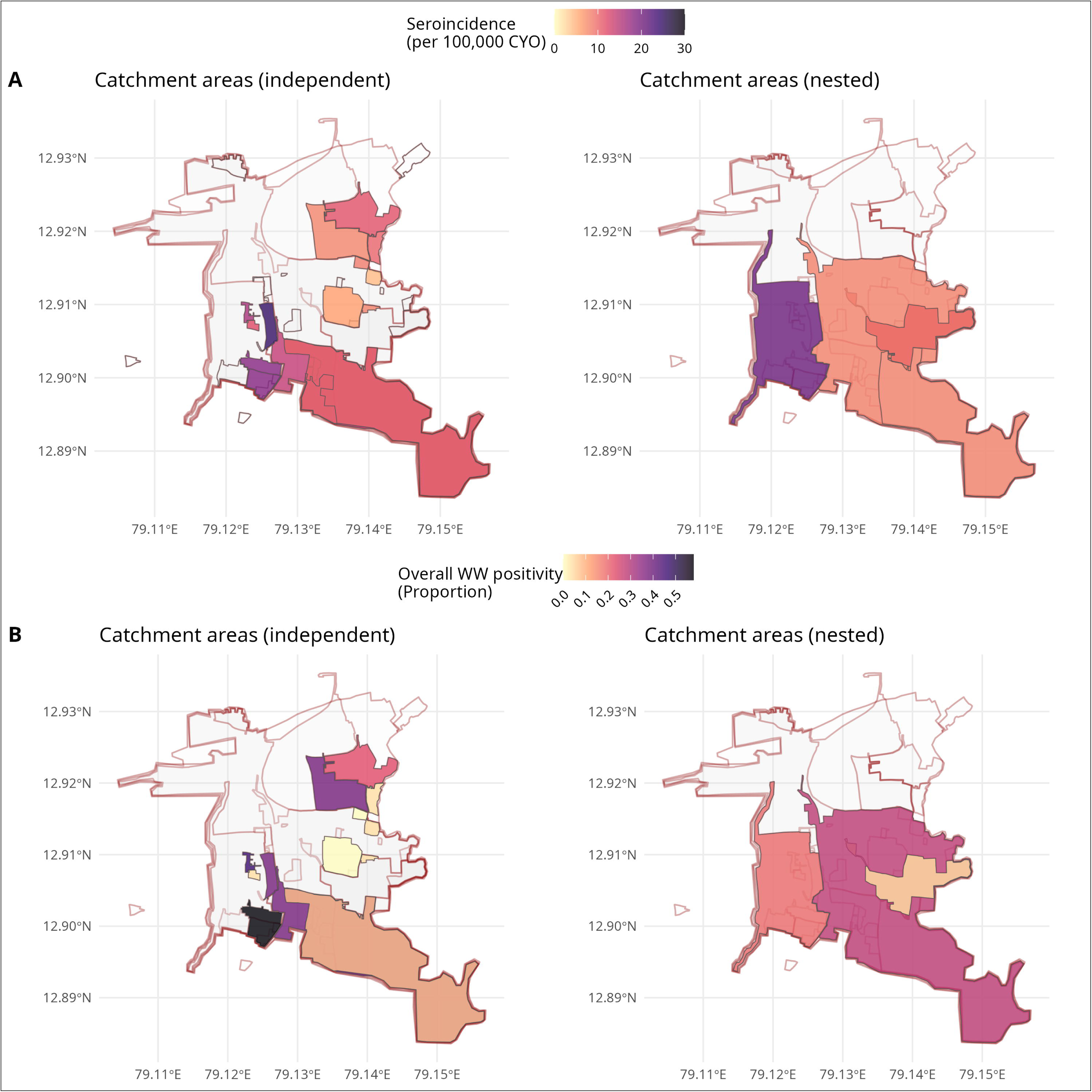
Seroincidence of typhoid and S. Typhi positivity in wastewater across independent and nested catchments. Maps illustrate the 20 catchment areas (out of 40) included in the serosurvey. Panel A shows both independent catchment areas and larger overlapping catchments, with the gradient of the coloured polygons representing the estimated seroincidence. Panel B shows the coloured polygons indicating overall wastewater positivity across the catchment areas.

### Association of seroincidence with detection of *S.* Typhi in wastewater

Negative binomial regression analysis was performed to account for overdispersion in the data to estimate the association between seroincidence and overall WW positivity, adjusting for the population size of the catchment areas. The model for grab sample positivity as a response to seroincidence with the catchment population as an additional predictor provided the best fit (LLR = −29.11). *S.* Typhi detections in WW positively correlated with seroincidence (incidence rate ratio (IRR) 1.14 (95% CI: 1.07–1.23, *p* < 0.001) for grab samples and 1.10 (95% CI: 1.02–1.20, *p* = 0.032) for Moore swabs; Table 2). The upstream catchment population was associated with the detection of *S*. Typhi; however, this association became non-significant when seroincidence was considered (Table 2).

**Table 2.**
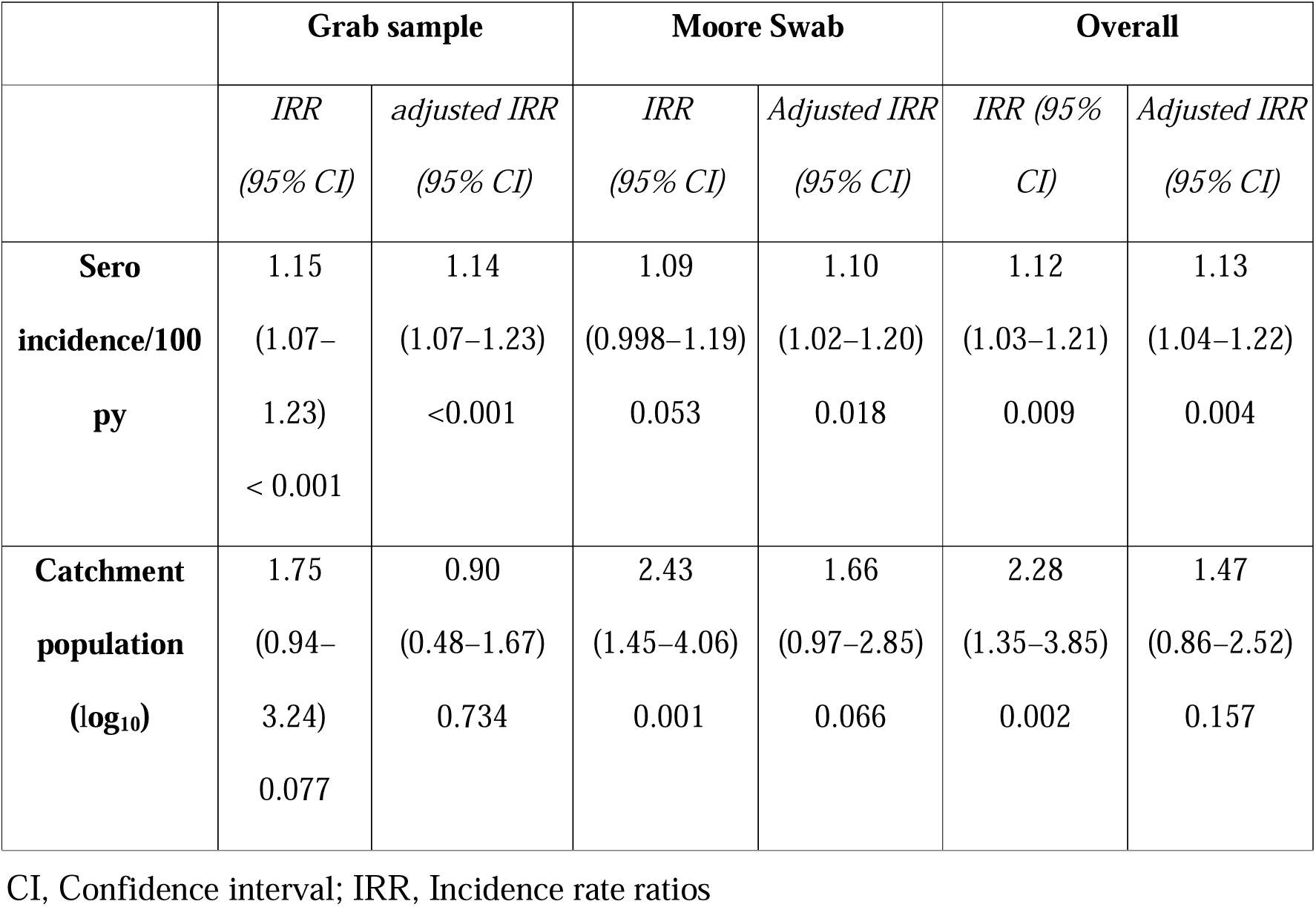
Typhoid seroincidence predicting ES positivity.

## Discussion

This study originally aimed to demonstrate the effectiveness of ES of WW as an alternative to blood culture-based surveillance for quantifying typhoid burden in resource-limited settings. Due to a lack of clinical data, we could not demonstrate this, but we found that ES of WW for *S.* Typhi in urban settings of low-middle-income contexts is implementable. In an earlier case-control study, environmental samples from households of participants diagnosed with typhoid were processed using both grab sampling and Moore swabs [32]. The odds ratio (OR) of WW positivity from typhoid-positive households was 1.34 for Moore swabs and 4.17 for grab sampling using the Bag-mediated filtration system (BMFS). However, the pandemic and ensuing lockdowns largely modified health-seeking behaviour, and the number of cases of blood culture-confirmed typhoid was approximately a tenth of what was seen during 2017 – 2020 in the same region. The current study aimed to leverage ongoing clinical surveillance for typhoid to examine the association between community WW positivity and disease burden using a detailed sampling scheme and systematic, repeated measurements of *S.* Typhi detection from sites with a defined upstream catchment.

The positivity of Moore swabs (15.30%) was two-fold higher than that of grab samples (7.50%). This finding aligns with the findings of a previous study [32], where grab sampling using the BMFS method showed 3.42% positivity, and Moore swabs showed 8.67%. Grab samples reflect the *S.* Typhi status in WW at collection time, while Moore swabs capture particulates and organisms during deployment. Moore swab data, inherently non-quantitative, are more suitable when detection sensitivity is crucial, such as estimating disease burden post-public health intervention. During outbreaks, increased genome copy numbers can signal above-expected endemic or seasonal levels. Studies indicate Moore swab sensitivity inversely correlates with sewer size and directly correlates with the number of swabs deployed [17,33]. Therefore, sample processing methods should be chosen based on specific use cases.

The seasonality of blood culture-confirmed enteric fever was indiscernible due to limited clinical case data. Previously, the SEFI study had identified variations in clinical case detection over three years in Vellore, with no correlation to the monsoon months [4]. Another report from the same area noted a typhoid outbreak from April to June 2019 during the summer [34]. A global review [35] found that seasonality in the occurrence of typhoid cases and its association with rainfall was more pronounced farther from the equator, with peaks from May and October in Asia. However, this review included only one study from India, indicating the need for more comprehensive and systematic research on typhoid seasonality in the country.

The present study revealed a notable increase in WW positivity trends before the wetter months for both grab and Moore swabs. The effect of variations in the physicochemical characteristics of WW and molecular detection has been established, and the Centers for Disease Control and Prevention recommends matrix recovery controls and fecal normalization for WW surveillance for SARS-CoV-2 [36], but the actual association with positivity remains unclear. In the present study, significant between- and within-site variations were observed in these characteristics, but they did not significantly correlate with the detection of *S.* Typhi in WW grab samples. This may be due to the inherently low levels of *S.* Typhi in WW, contrasting with viruses like SARS-CoV-2, which show a clearer association between WW characteristics and positivity.

The study revealed a significant association between detection and upstream catchment population for both grab and Moore swab sampling, with a stronger association for Moore swab positivity than that for grab sampling. This was anticipated since larger populations exhibit higher typhoid event rates. Continuous filtration through Moore swab sampling is more pertinent for estimating typhoid burden when sensitivity is crucial rather than quantifying shedding during outbreaks or high incidence periods [10].

Normalization of WW pathogen detection by levels of fecal contamination must account for variations in catchment populations, sampling networks, and dilution factors like rainfall and flow rates. HF183 levels, indicating fecal contamination, correlated with *S.* Typhi detection in Moore swabs but not in grab samples. We found no correlation between HF183 levels and upstream catchment population or rainfall. This lack of association in our setting may result from dilution effects, where higher fecal contamination in densely populated areas is diluted by increased sewage flow, leaving HF183 levels unaffected by upstream population, or a saturation effect, where qPCR is inhibited by high *Bacteroides* spp. levels, yielding similar values above a threshold. A recent study also showed HF183 levels did not correlate with population density, nor was detection frequency affected by rainfall [37]. Together, these studies indicate that alternative markers of human fecal contamination, such as CrAssphage or pepper mild mottle virus, which correlate with flow and catchment populations, should be considered.

During the SARS-CoV-2 pandemic, community lockdowns restricted passive hospital-based surveillance, affecting clinical incidence estimates and healthcare-seeking behaviours. The annual seroincidence of typhoid from the survey was 10.40/100 py, approximately ten times higher than the incidence of blood culture-confirmed disease reported in the SEFI study. This discrepancy may result from seroincidence capturing both subclinical and clinical cases, suggesting that seroincidence could more effectively measure vaccine impacts on typhoid transmission. A previous study estimated seroincidence rates from cross-sectional surveys of children in four regions and reported seroincidence rates of 17.60/100 py in Karachi, Pakistan and 41.20/100 py in Dhaka, Bangladesh [18]. Furthermore, in Nepal the study also reported that while the clinical incidence of typhoid fever increased with age, seroincidence did not, likely due to multiple exposures and altered immune decay in different age groups. Therefore, further studies using this surveillance method are required for further comparison. Seroincidence varied across catchment areas, suggesting potential transmission hotspots and emphasizing the need for selecting appropriately sized populations for surveillance studies.

WW positivity via grab sampling exhibited a median membrane filtration positivity of 7.69%, irrespective of catchment size. Moore swab positivity was influenced by upstream catchment size, showing median positivity of 7.69% in non-nested catchments, 15.40% and 38.46% in level 1 and larger catchments respectively. Similar to HF183 detection, this may be due to a dilution effect, as grab sample positivity is balanced by increased flow at larger catchments, whereas Moore swabs collect more bacteria due to extended sewage exposure. The correlation between seroincidence and grab sample positivity was stronger than that between Moore swab positivity, even though Moore swabs showed twice the positivity. These associations remained unaffected after controlling for the catchment populations in the model.

This study is among the first to estimate the association between ES positivity for *S.* Typhi and seroincidence of typhoid in the community. The feasibility of WW surveillance in LMICs has been shown for SARS-CoV-2 [38,39]. We aimed to standardize methods for detecting *S.* Typhi in WW and demonstrated the feasibility of scaling up ES. Clinical surveillance for illnesses with low incidence and nonspecific symptoms often underestimates incidence due to missed asymptomatic infections and carriers. The methods discussed— cross-sectional serosurveys and ES—offer alternative strategies for estimating burden in resource-limited settings where intensive clinical surveillance is impractical.

This study has several limitations. WW is categorized as positive for S. Typhi when PCR has detected all three molecular targets, however, this assumption can be violated if there are strains of bacteria that can give non-specific results for one or more of these targets if they are present in the right combination. The clinical incidence estimation is constrained by altered healthcare-seeking behaviors in the community due to the COVID-19 pandemic, which, along with lockdowns, may have also altered typhoid transmission dynamics. The HlyE IgG is not specific to *S*. Typhi and can also be increased by *S.* Paratyphi A infection, which has not been assessed in the present study. Serosurvey was not carried out in all catchments of the study area but only in half the catchments since a comparative analysis was not initially planned. Nonalignment in WW sampling and sero-survey timing could have affected the study findings. Seroincidence estimation assumes that antibody decay rates remain constant for a group while asymptomatic infection or reinfection may modify antibody responses. Long-term carriage may potentially alter immune responses; however, a 2013 study conducted immunoscreening in chronic Salmonella Typhi carriers did not identify HlyE as a reactive antigen [40].

## Conclusions

Despite the study’s limitations, including the focus on a single region and the need for more extensive sampling and longitudinal data, it provides valuable insights into using ES and seroincidence surveys for monitoring and responding to typhoid outbreaks. Our findings indicate that ES could complement traditional surveillance methods effectively, providing valuable insights for public health interventions and disease monitoring in resource-limited settings. Future research should aim to expand these methodologies across diverse geographic and seasonal contexts to strengthen their applicability and effectiveness in typhoid control efforts.

## Data Availability

All data produced are available online at https://github.com/drvenkatm/ES_Salmonella_Vellore-May22_Apr23.git

## Acknowledgements

We want to acknowledge Annai Gunasekaran and Senthil Kumar for their contributions in coordinating the field components and Nirmal Kumar’s contribution to the laboratory work.

## Author Contributions

**Conceptualization**: Dilip Abraham, Christopher Uzzell, Nicholas Grassly, Venkata Raghava Mohan

**Methodology** - Dilip Abraham, Lalithambigai Kathiresan, Midhun Sasikumar, Kristen Aiemjoy, Richelle C. Charles, Rajan Srinivasan, Catherine Troman, Elizabeth Gray, Jacob John, Balaji Veeraraghavan, Nicholas Grassly, Venkata Raghava Mohan

**Formal Analysis**- Dilip Abraham, Lalithambigai Kathiresan, Dilesh Kumar, Venkata Raghava Mohan

**Validation** - Dilip Abraham, Lalithambigai Kathiresan, Nicholas Grassly, Venkata Raghava Mohan

**Writing** – original draft - Dilip Abraham, Lalithambigai Kathiresan, Midhun Sasikumar, Venkata Raghava Mohan

**Writing – review & editing** - Dilip Abraham, Lalithambigai Kathiresan, Midhun

Sasikumar, Kristen Aiemjoy, Richelle C. Charles, Dilesh Kumar, Rajan Srinivasan, Catherine Troman, Elizabeth Gray, Christopher Uzzell, Jacob John, Balaji Veeraraghavan, Nicholas Grassly, Venkata Raghava Mohan

