## Supplementary tables for "Wastewater surveillance for *Salmonella* Typhi and its association with seroincidence of enteric fever in Vellore, India"

**Supporting information**

**S1 Table.** Physico-chemical characteristics of wastewater – between-site and within-site variation.

| ***Variable*** | ***Median (IQR)*** | ***Between site variation*** | | ***Within-site variation (repeated)*** | |
| --- | --- | --- | --- | --- | --- |
|  |  | ***F Statistic*** | ***p Value*** | ***F Statistic*** | ***p Value*** |
| Temperature (ºC) | 27.8 (26.3 - 28.9) | 0.97 | 0.518 | 20.66 | < 0.001 |
| pH | 7.2 (7.08 - 7.43) | 1.63 | 0.011 | 6.22 | 0.013 |
| Oxidative reductive potential (mV) | -234.5 (-299.5 to -150.6) | 5.17 | <0.001 | 23.71 | < 0.001 |
| Dissolved oxygen (%saturation) | 39.8 (28.9 - 51.1) | 3.07 | <0.001 | 27.63 | < 0.001 |
| Total dissolved solids (mg/L) | 582.5 (452 -754) | 1.87 | <0.001 | 66.59 | < 0.001 |
| Turbidity (NTU) | 107.5 (67.7 - 160) | 5.68 | <0.001 | 8.70 | 0.003 |

IQR – interquartile range; mV – millivolt; mg/L – milligrams/litre, NTU – nephalometric turbidity unit

**S2 Table.** Serosurvey participants and seroincidence by catchment areas of sampling sites.

| ***Location ID*** | ***Catchment stratification*** | ***Number of serosurvey participants*** | ***Seroincidence/ 100py*** | ***95% CI*** |
| --- | --- | --- | --- | --- |
| 106 | 1 | 45 | 18.7 | 12.08 – 28.90 |
| 108 | 1 | 125 | 14.3 | 11.19 – 18.40 |
| 110 | 1 | 34 | 24.6 | 15.47 – 39.22 |
| 111 | 1 | 23 | 16.1 | 9.30 – 27.74 |
| 112 | 1 | 31 | 12 | 6.81 – 21.03 |
| 120 | 1 | 20 | 8.6 | 4.15 – 17.79 |
| 122 | 1 | 216 | 10.3 | 8.36 – 12.77 |
| 123 | 1 | 82 | 4.2 | 2.64 – 6.72 |
| 127 | 1 | 27 | 8.2 | 4.14 – 16.37 |
| 130 | 1 | 71 | 6.1 | 3.88 – 9.69 |
| 137 | 1 | 53 | 10.4 | 6.93 – 15.51 |
| 139 | 1 | 210 | 11.6 | 9.44 – 14.18 |
| 143 | 1 | 61 | 7.9 | 5.28 – 11.74 |
| 114 | 2 | 103 | 9.5 | 6.98 – 12.82 |
| 116 | 2 | 494 | 8.3 | 7.13 – 9.56 |
| 145 | 2 | 184 | 20.6 | 16.82 – 25.31 |
| 149 | 2 | 101 | 9.7 | 7.15 – 13.13 |
| 140 | 3 | 887 | 10.3 | 9.25 – 11.36 |
| 141 | 3 | 725 | 10.2 | 9.13 – 11.46 |
| 142 | 3 | 1172 | 10.5 | 9.56 – 11.43 |

**S1 Fig.** Wastewater positivity and seroincidence in catchment areas

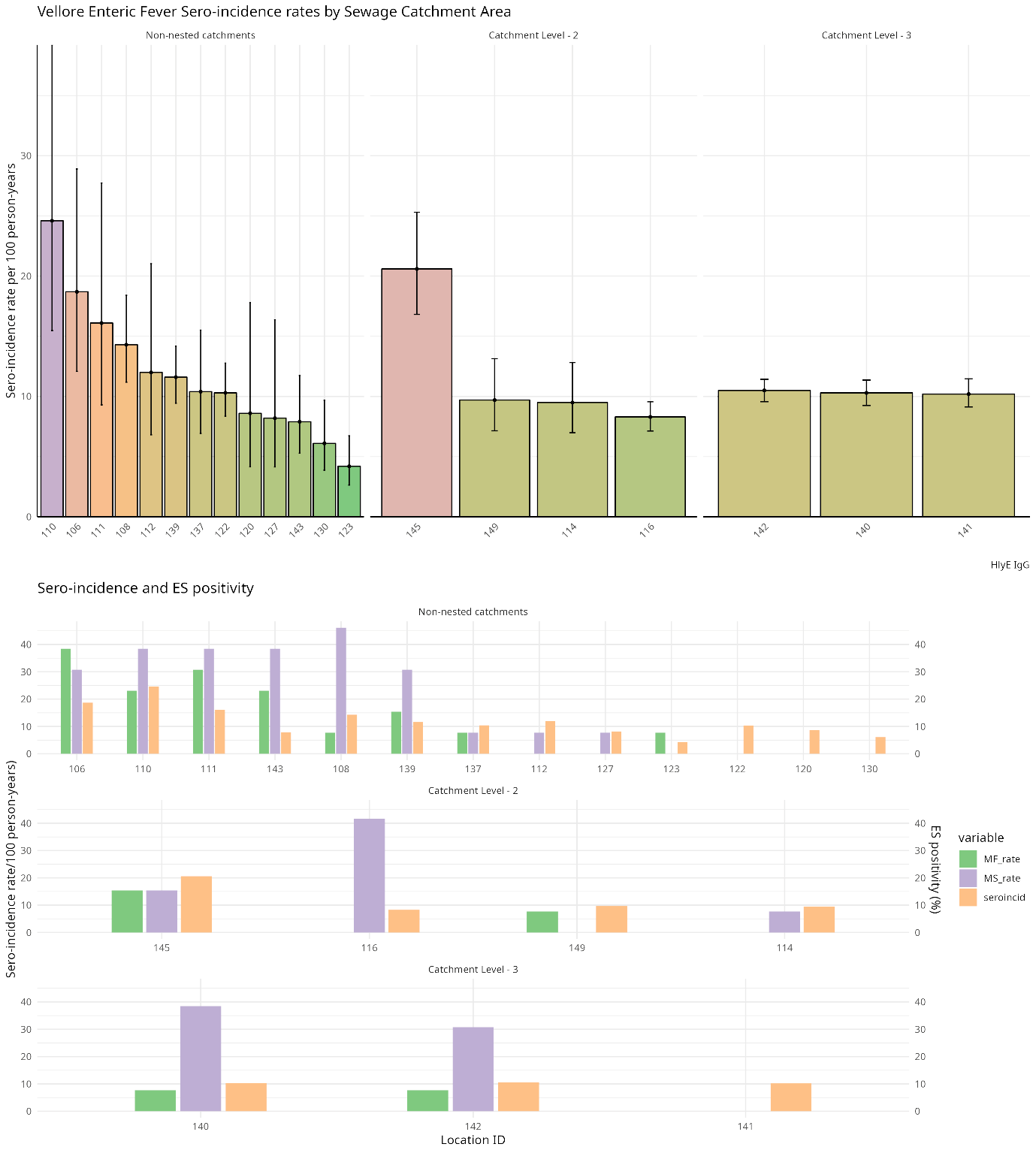
